# Diagnostic Value of Symptoms for Pediatric SARS-CoV-2 Infection in a Primary Care Setting

**DOI:** 10.1101/2021.03.29.21254600

**Authors:** Chien-Hsiang Weng, Wesley Wing Wah Butt, Meredith B. Brooks, Claudia Clarke, Helen E. Jenkins, Sabina D. Holland, Silvia S. Chiang

## Abstract

**Purpose:** To evaluate the diagnostic value of symptoms used in the screening approaches by daycares and schools for identifying children and adolescents with possible SARS-CoV-2 infection, we designed a large observational study utilizing the data from primary care settings.

**Methods:** This cohort study included children and adolescents evaluated in a network of clinics in Rhode Island. Participants were age-stratified: 0-4, 5-11, and 12-17 years. We estimated the sensitivity, specificity, and area under the receiver operating curve (AUC) of individual symptoms and three symptom combinations: a probable case definition published by the Rhode Island Department of Health (RIDOH), and two novel combinations generated by different statistical approaches to maximize sensitivity and AUC. We evaluated the test characteristics of symptom combinations both with and without consideration of COVID-19 exposure.

**Results:** Two-hundred seventeen (39.1%) of 555 participants were SARS-CoV-2-infected. Fever was more common among 0-4 years-olds (p=0.002); older children more frequently reported fatigue (p=0.02) and anosmia or ageusia (p=0.047). In children >5 years old, anosmia or ageusia had 94-98% specificity. In all age groups, exposure history most accurately predicted infection. In combination with COVID-19 exposure history, various symptom combinations had sensitivity >95% but specificity <30%. No individual symptom or symptom combination had AUC ≥0.70.

**Conclusions:** Anosmia or ageusia in children ≥5 years old and dyspnea in children 5-11 years old should raise providers’ index of suspicion for COVID-19. However, our overall findings underscore the limited diagnostic value of symptoms and the critical need for widely available, efficient testing.

## Introduction

Severe acute respiratory syndrome coronavirus 2 (SARS-CoV-2) has caused coronavirus disease 2019 (COVID-19) in 30 million people in the United States (U.S.) as of late March 2021.(1) Children and adolescents account for 12.3% of reported COVID-19 cases in the U.S.(2) Most children and adolescents have mild symptoms and are managed as outpatients(3–5); however, few clinical studies of pediatric COVID-19 have been conducted in primary care settings.(6–8) A clearer understanding of the diagnostic value of symptoms may have implications for the symptom screening approaches used in daycares and schools for identifying children and adolescents with possible SARS-CoV-2 infection.

To estimate the accuracy of symptoms for identifying pediatric SARS-CoV-2 infection, we conducted this study of children and adolescents evaluated for COVID-19 in a network of clinics in Providence, Rhode Island. We assessed the test characteristics of individual symptoms and three symptom combinations: the Rhode Island Department of Health (RIDOH) probable case definition,(9) which is used to screen students and daycare attendees for COVID-19, and two novel combinations generated by statistical approaches to maximize sensitivity and area under the receiver operating curve (AUC). As a secondary aim, we evaluated demographic and clinical predictors of SARS-CoV-2 infection in our pediatric patients.

## Methods

### Setting

This retrospective cohort study took place between March 20-June 22, 2020 before the massive asymptomatic screening were recommended, at the Providence Community Health Centers (PCHC), a network of ten clinics that provide primary care, urgent care, and specialty services. PCHC serves approximately 60,000 patients, who are predominantly Hispanic. Ninety percent of patients have household incomes under 200% of the federal poverty level.(10) Before the study start date, PCHC clinicians implemented a standardized template to document symptoms and exposure history of patients under evaluation for COVID-19. On April 1, 2020, RIDOH recommended SARS-CoV-2 testing for anyone with exposure to or symptoms of COVID-19.(11) Concurrently, local hospitals implemented pre-admission and pre-procedure COVID-19 screening.

### Participants

We identified all PCHC patients who received SARS-CoV-2 reverse transcriptase polymerase chain reaction (RT-PCR) testing on a nasopharyngeal sample on or before June 22, 2020, and were younger than 18 years old at the time of the test. We included patients whose exposure and symptoms were evaluated either before or after PCR testing. The latter group consisted of patients tested in an emergency department or who underwent pre-procedure or pre-admission COVID-19 screening, as long as their PCR result was documented in their PCHC chart and they were evaluated with the standardized template during a follow-up visit.

### Data collection and variables

Three authors (CW, WB, CC) manually abstracted the following variables: age, sex, self-reported race/ethnicity, insurance status, body mass index (BMI)-for-age percentile, history of asthma or allergic rhinitis, COVID-19 exposure history, presenting symptoms, and type of encounter that led to SARS-CoV-2 testing (PCHC primary care, PCHC urgent care, emergency department, procedure, or hospital admission). On the standardized template, the following symptoms were marked as present or absent: new cough, dyspnea, new congestion/rhinorrhea, myalgia, fever ≥100.4°F, headache, sore throat, abdominal pain, nausea, vomiting, diarrhea, anosmia or ageusia, and fatigue.

We categorized participants into age groups corresponding roughly with U.S. educational stages: 0-4 years (daycare/preschool), 5-11 years (elementary school), and 12-17 years (middle/high school). Race/ethnicity was grouped into Hispanic, non-Hispanic (NH) Black, NH White, and NH other (Asians, other Pacific Islanders, more than one race, and unknown). We used CDC definitions to categorize BMI-for-age percentile.(12) Known COVID-19 exposure was self-reported and defined as contact with a confirmed or suspected COVID-19 patient ≤14 days prior to SARS-CoV-2 testing.

### Statistical analysis

We performed univariable binomial regression to compare demographic and clinical characteristics between SARS-CoV-2-infected and uninfected participants. Variables that differed between the two groups at a significance level of p<0.2 were retained in a multivariable model. We checked for interactions on the multiplicative scale between known COVID-19 exposure and all other covariates.

For each age group, we calculated the sensitivity, specificity, and AUC of COVID-19 exposure history and each symptom for identifying SARS-CoV-2 infection. Myalgia, headache, sore throat, abdominal pain, nausea, and anosmia or ageusia were not assessed in children 0-4 years old due to their lower ability to report these symptoms. We then evaluated the diagnostic value of three symptom combinations: (1) the probable case definition published by RIDOH(9); (2) a combination generated by a backward elimination approach; and (3) a combination generated by classification and regression tree (CART) analysis. We evaluated the test characteristics of the three symptom combinations both with and without consideration of COVID-19 exposure.

According to the RIDOH criteria, a probable COVID-19 case has one of the following: new cough, shortness of breath, or anosmia or ageusia. A case also qualifies as probable COVID-19 by having at least two of the following: fever, chills, myalgia, headache, sore throat, nausea or vomiting, diarrhea, fatigue, or new congestion or rhinorrhea.(9) As we did not collect information on chills, we excluded this symptom in our analysis of the RIDOH criteria.

For each age group, we used a backward elimination approach to generate a symptom combination that maximized specificity without sacrificing sensitivity. First, we calculated the sensitivity, specificity, and AUC if any of the symptoms were present (the baseline combination). Then, we manually removed symptoms one at a time, in order of ascending AUC. Symptoms with the same AUC were removed in order of ascending sensitivity. We selected the combination with the highest specificity but the same sensitivity as the baseline combination.

We used CART analysis to identify the symptoms that best predicted SARS-CoV-2 infection in each age group. In CART analysis, measures of predictive importance were assigned to each symptom, entailing both marginal and interaction effects involving this variable. The data set was then split into increasingly homogenous sub-groups, using improvement in the Gini gain score, to identify the explanatory variable that gave the best discrimination between the two outcome classes (COVID-19 vs. no COVID-19). Maximal trees were created and then pruned based on relative misclassification costs, complexity, and parsimony. Ten-fold cross-validation was performed, in which the whole data set was randomly split into learning and test data sets. CART analysis was then applied to determine model performance and predictive accuracy in these test sets, removing the need for a validation data set. We calculated discriminatory properties of having at least one of the most important symptoms identified as nodes on the final derived trees for each age group.

To determine the impact of recall bias from applying the standardized template after PCR testing, we performed a sensitivity analysis restricted to participants who were evaluated for exposure and symptoms before testing. We decided *a priori* to assess the diagnostic value of symptom combinations only if there were meaningful differences in AUCs of exposure history and individual symptoms.

Twenty-two (4.0%) participants with unknown race/ethnicity were grouped into the “NH other” group and included in all analyses. BMI-for-age percentile—which is measured in children at least two years old—was missing for 128 (23.1%) participants, 96 of whom were younger than two years. These participants were excluded from comparisons of BMI between SARS-CoV-2-infected and uninfected children only. Four (0.7%) participants had no data for one symptom; they were excluded from regression models that examined the association between the number of presenting symptoms and SARS-CoV-2 infection, as well as from calculations of sensitivity, specificity, and AUC for the missing symptom only. There were no other missing data.

Analyses were conducted using R version 3.5.1 (R Statistical Computing, Vienna, Austria) and Salford Systems Data Mining and Predictive Analytics Software version 8.0 (Salford Systems, San Diego, California, U.S.). Sensitivity, specificity, and AUC estimates were calculated with the reportROC package for R.(13)

### Ethics

The PCHC Human Subjects Review Committee approved this study and waived informed consent.

## Results

Before June 22, 2020, SARS-CoV-2 PCR was performed in 803 individuals <18 years of age who were registered as PCHC patients. We included 555 (69.1%) who were evaluated using the standardized template. PCHC clinicians assessed 506 (91.2%) participants in primary care and five (0.9%) in urgent care prior to PCR testing. Forty-four (7.9%) participants were assessed by PCHC clinicians after pre-procedure screening (n=10), hospital admission (n=21), and emergency room visit (n=13). The 248 excluded patients were tested at a PCHC specialty clinic or a non-PCHC facility without subsequent evaluation using the standardized template (Supplementary Material).

Of the 555 participants, 283 (51.0%) had known COVID-19 exposure and at least one symptom; 183 (33.0%) had at least one symptom but no known exposure; 56 (10.1%) had known exposure but no symptoms; and 33 (5.9%) participants had neither symptoms nor known exposure. Two-hundred eighty-nine (52.1%) participants were tested after reopening on May 9. Median age was 9 (IQR: 3.5-15) years; 293 (52.8%) were female, 459 (82.7%) were Hispanic, and 37 (6.7%) were uninsured (Table 1).

**Table 1:** Patient Characteristics at Time of SARS-CoV-2 reverse transcriptase polymerase chain reaction

|  | n (%) or median (IQR) |  |  | Univariable regression |  | Multivariable regression |  |
| --- | --- | --- | --- | --- | --- | --- | --- |
|  | Uninfected<br>(n=338) | Infected<br>(n=217) | All<br>(n=555) | OR (95% CI) | p-value | aOR (95% CI) | p-value |
| Age | 8 (3-14) | 11 (6-15) | 9 (3.5-15) | 1.06 (1.03, 1.09) | <0.001 | 1.00 (0.97, 1.04) | 0.88 |
| Female | 183 (54.1) | 110 (50.7) | 293 (52.8) | 0.87 (0.62, 1.23) | 0.43 |  |  |
| Race/ethnicity<sup>a</sup> |  |  |  |  | <0.001 |  |  |
| Hispanic | 257 (76.0) | 202 (93.1) | 459 (82.7) | 3.30 (1.32, 10.02) | 0.018 | 3.73 (1.36, 12.10) | 0.016 |
| NH Black | 21 (6.2) | 5 (2.3) | 26 (4.7) | Ref |  | Ref |  |
| NH White | 24 (7.1) | 6 (2.8) | 30 (5.4) | 1.05 (0.28, 4.12) | 0.94 | 1.91 (0.43, 8.78) | 0.40 |
| NH Other | 36 (10.7) | 4 (1.8) | 40 (7.2) | 0.47 (0.11, 1.95) | 0.29 | 0.60 (0.14, 2.75) | 0.51 |
| Uninsured | 17 (5.0) | 20 (9.2) | 37 (6.7) | 1.92 (0.98, 3.79) | 0.054 | 1.68 (0.76, 3.81) | 0.20 |
| BMI category<sup>b</sup> |  |  |  |  | 0.20 |  |  |
| Underweight | 9 (3.6) | 2 (1.1) | 11 (2.6) | 0.36 (0.05, 1.43) |  |  |  |
| Normal | 127 (50.2) | 79 (45.4) | 206 (48.2) | Ref |  |  |  |
| Overweight | 40 (15.8) | 37 (21.3) | 77 (18.0) | 1.49 (0.88, 2.52) |  |  |  |
| Obese | 77 (30.4) | 56 (32.2) | 133 (31.1) | 1.17 (0.75, 1.82) |  |  |  |
| Asthma or allergic rhinitis | 70 (20.7) | 40 (18.4) | 110 (19.8) | 0.87 (0.56, 1.33) | 0.51 |  |  |
| Known COVID-19 exposure | 150 (44.4) | 189 (87.1) | 339 (61.1) | 8.46 (5.46, 13.51) | <0.001 | 8.75 (5.47, 14.46) | <0.001 |
| Number of COVID-19 symptoms | 2 (1-3) | 3 (2-4) | 2 (1-4) | 1.32 (1.20, 1.46) | <0.001 | 1.32 (1.18, 1.47) | <0.001 |
| Timing of PCR |  |  |  |  |  |  |  |
| During stay-at-home order | 158 (59.4) | 108 (40.6) | 266 (47.9) | Ref |  |  |  |
| After reopening | 180 (62.3) | 109 (37.7) | 289 (52.1) | 0.89 (0.63, 1.25) | 0.49 |  |  |

<sup>a</sup>Of 459 Hispanics, 2 were Asian; 13, Black; 247, White, 85, >1 race; 3, Pacific Islander; 109, Unknown. Of 40 “Other,” 9 were Asian; 8, >1 race; 1, Pacific Islander; 22, Unknown.
<sup>b</sup>BMI category applies only to children age $\geq 2$ years. In addition, 18 uninfected and 14 infected children $\geq 2$ years were missing BMI category; therefore, the percentages for the uninfected group and the infected group are calculated from 254 and 174, respectively.
Abbreviations: BMI, body mass index; aOR, adjusted odds ratio; CI, confidence interval; IQR, interquartile range; OR, odds ratio; Ref, reference category.
Variables that were not included in the adjusted model have empty cells in the multivariable regression.

Two-hundred seventeen (39.1%) participants had SARS-CoV-2 infection. One infected participant had neither known exposure nor symptoms. Asymptomatic infections occurred in 2/40 (5.0%) 0-4 year-olds, 9/69 (13.0%) 5-11 year-olds, and 9/108 (8.3%) 12-17 year-olds (Figure 1). Twenty-eight (12.9%) SARS-CoV-2-infected participants had only one symptom (Table 2).

**Table 2:** Participants Presenting with Only One Symptom

|  | No. (%) participants |  |
| --- | --- | --- |
|  | Uninfected (n=72) | Infected (n=28) |
| Symptom |  |  |
| Fever | 22 (30.6) | 11 (33.3) |
| Fatigue | 2 (2.8) | 0 (0.0) |
| Myalgia | 5 (6.9) | 2 (7.1) |
| Headache | 5 (6.9) | 3 (10.7) |
| Cough | 11 (15.3) | 6 (21.4) |
| Dyspnea | 3 (4.2) | 1 (3.6) |
| Sore throat | 3 (4.2) | 3 (10.7) |
| Congestion/rhinorrhea | 9 (12.5) | 1 (3.6) |
| Anosmia or ageusia | 2 (2.8) | 0 (0.0) |
| Abdominal pain | 1 (1.4) | 0 (0.0) |
| Nausea/vomiting | 4 (5.6) | 0 (0.0) |
| Diarrhea | 5 (6.9) | 1 (3.6) |
| Known COVID-19 exposure | 33 (45.8) | 24 (85.7) |

**Figure 1:**
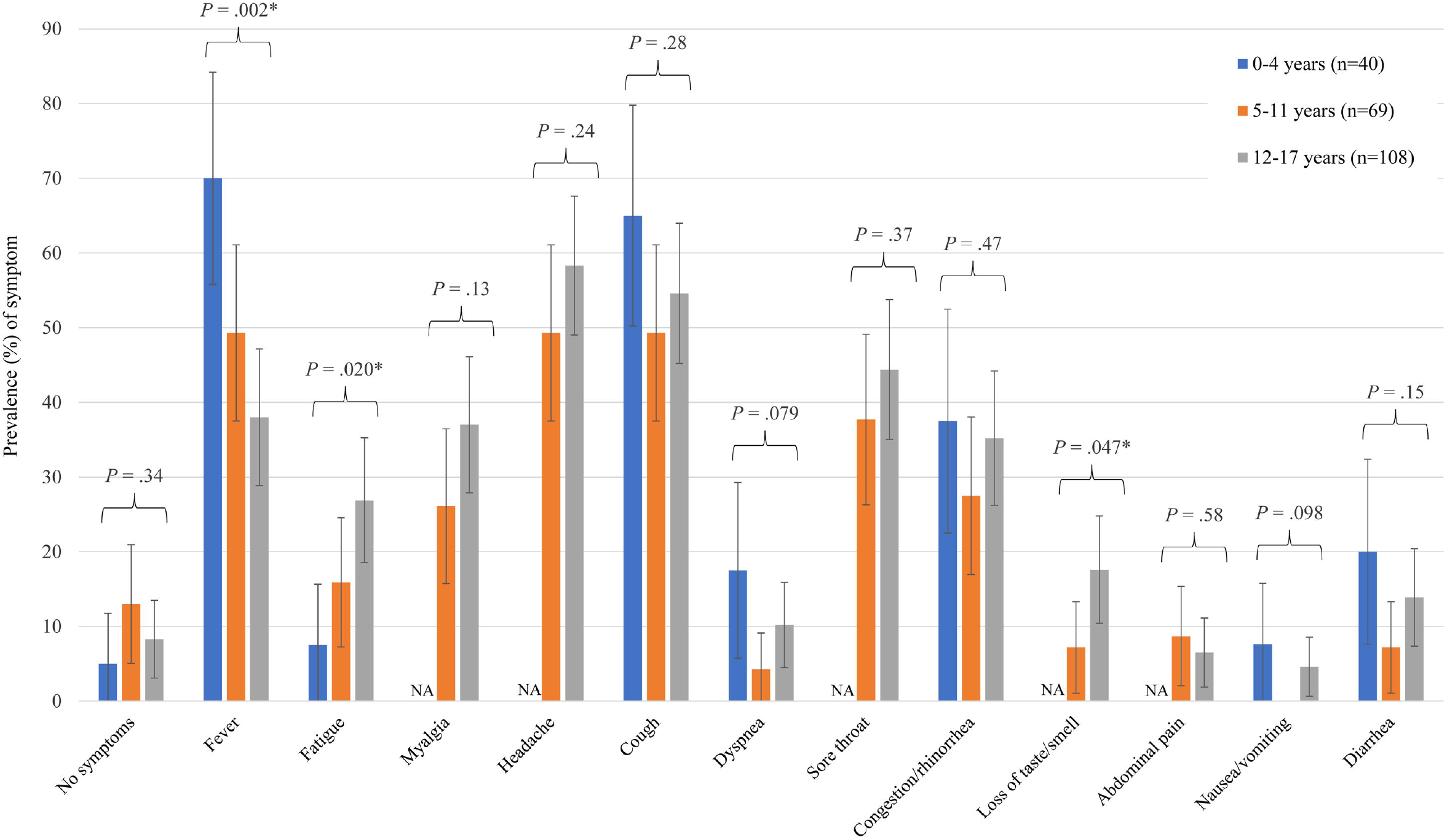
Age-Stratified Clinical Presentation among 217 SARS-CoV-2-Positive Participants. Myalgia, headache, sore throat, abdominal pain, nausea, and loss of taste/smell were not assessed in children 0-4 years-old due to the lower reliability of these symptoms in this group. Bars show 95% confidence intervals. No children between the ages of 5-11 years presented with nausea/vomiting. Abbreviation: NA, not applicable.

Children with a positive PCR were more likely to be older (11 vs. 8 years), have known COVID-19 exposure (87.1 vs 44.4%), be Hispanic (93.1 vs. 76.0%), and present with more symptoms (3 vs. 2 symptoms). Test positivity did not differ between participants evaluated before and after reopening. The multivariable regression analyses showed consistent results (Table 1).

Stratifying the 217 children with COVID-19 by age, we observed fever more frequently among children aged 0-4 years (p=0.002). The prevalence of fatigue increased with age (p=0.02). Adolescents 12-17 years old (p=0.047) were more likely to present with anosmia or ageusia compared to children aged 5-11 years (Figure 1; Supplementary Material).

In all age groups, known COVID-19 exposure alone had the highest AUC for identifying SARS-CoV-2 infection. No individual symptom or symptom combination had AUC >0.7 (Tables 3–5). When exposure history was considered, all symptom combinations had 97-100% sensitivity.

**Table 3:** Diagnostic Value of Symptom Screening in Children 0-4 Years of Age

|  | No. (%) participants with symptom |  | p-value | Sensitivity<br>(95% CI) | Specificity<br>(95% CI) | AUC |
| --- | --- | --- | --- | --- | --- | --- |
|  | Uninfected<br>(n=115) | Infected<br>(n=40) |  |  |  |  |
| Known COVID-19 exposure | 31 (27.2) | 31 (77.5) | <0.001 | 77.5 (64.6-90.4) | 72.8 (64.6-81.0) | 0.75 |
| *Individual symptoms* |  |  |  |  |  |  |
| Cough | 31 (27.2) | 26 (65.0) | <0.001 | 65.0 (50.2-79.8) | 72.8 (64.6-81.0) | 0.69 |
| Fever | 61 (53.5) | 28 (70.0) | 0.069 | 70.0 (55.8-84.2) | 46.5 (37.3-55.6) | 0.58 |
| Congestion/rhinorrhea | 25 (21.9) | 15 (37.5) | 0.053 | 37.5 (22.5-52.5) | 78.1 (70.5-85.7) | 0.58 |
| Dyspnea | 13 (11.4) | 7 (17.5) | 0.32 | 17.5 (5.7-29.3) | 88.6 (82.8-94.4) | 0.53 |
| Diarrhea | 19 (16.7) | 8 (20.0) | 0.63 | 20.0 (7.6-32.4) | 83.3 (76.5-90.2) | 0.52 |
| Fatigue | 7 (6.1) | 3 (7.5) | 0.76 | 7.5 (0.0-15.7) | 93.9 (89.5-98.3) | 0.51 |
| Vomiting<sup>a</sup> | 15 (13.2) | 3 (7.7) | 0.36 | 7.7 (0.0-16.1) | 86.8 (80.6-93.0) | 0.47 |
| *Modified RIDOH probable case definition<sup>b</sup>* |  |  |  |  |  |  |
| Modified RIDOH<sup>c</sup> | 88 (77.2) | 38 (95.0) | 0.012 | 95.0 (88.2-100.0) | 22.8 (15.1-30.5) | 0.59 |
| Modified RIDOH<sup>c</sup> or known COVID-19 exposure | 99 (86.8) | 40 (100.0) | 0.016 | 100.0 (100.0-100.0) | 13.2 (7.0-19.4) | 0.57 |
| *Backward elimination<sup>b</sup>* |  |  |  |  |  |  |
| Cough, dyspnea, fever, congestion/rhinorrhea, or diarrhea | 87 (76.3) | 38 (95.0) | 0.009 | 95.0 (88.2-100.0) | 23.7 (15.9-31.5) | 0.59 |
| Cough, dyspnea, fever, congestion/rhinorrhea, diarrhea, or known COVID-19 exposure | 98 (86.0) | 40 (100.0) | 0.012 | 100.0 (100.0-100.0) | 14.0 (7.7-20.4) | 0.57 |

| *CART*<sup>b</sup> |  |  |  |  |  |  |
| --- | --- | --- | --- | --- | --- | --- |
| Cough | 31 (27.2) | 26 (65.0) | <0.001 | 65.0 (50.2-79.8) | 72.8 (64.6-81.0) | 0.69 |
| Cough, fever, or known COVID-19 exposure | 93 (81.6) | 39 (97.5) | 0.013 | 97.5 (92.7-100.0) | 18.4 (11.3-25.5) | 0.58 |

<sup>a</sup>There is a missing value for one participant.
<sup>b</sup>The participant was considered to be positive for the symptom combination if at least one of the features listed below was present.
<sup>c</sup>Cough (new) or shortness of breath alone, or two of any of the following: fever, fatigue, congestion/rhinorrhea (new), nausea/vomiting, and diarrhea.
Abbreviations: AUC, area under the receiver operating curve; CART, classification and regression tree; CI, confidence interval; RIDOH, Rhode Island Department of Health.

When exposure history was not considered, the RIDOH criteria and the combination generated by backward elimination had the highest sensitivity: 95% in 0-4 year-olds, 87% in 5-11 year-olds, and 92% in 12-17 year-olds. All combinations had <30% specificity.

In children <5 years old, fever and cough were the individual symptoms with the highest sensitivity at 70% and 65%, respectively (Table 3). In children 5-11 years old, no individual symptom had >50% sensitivity for COVID-19. Anosmia or ageusia had 98% specificity; dyspnea had 95% specificity (Table 4). Among adolescents 12-17 years old, cough and headache were the individual symptoms with the highest sensitivity. Anosmia or ageusia had 94% specificity (Table 5).

**Table 4:**
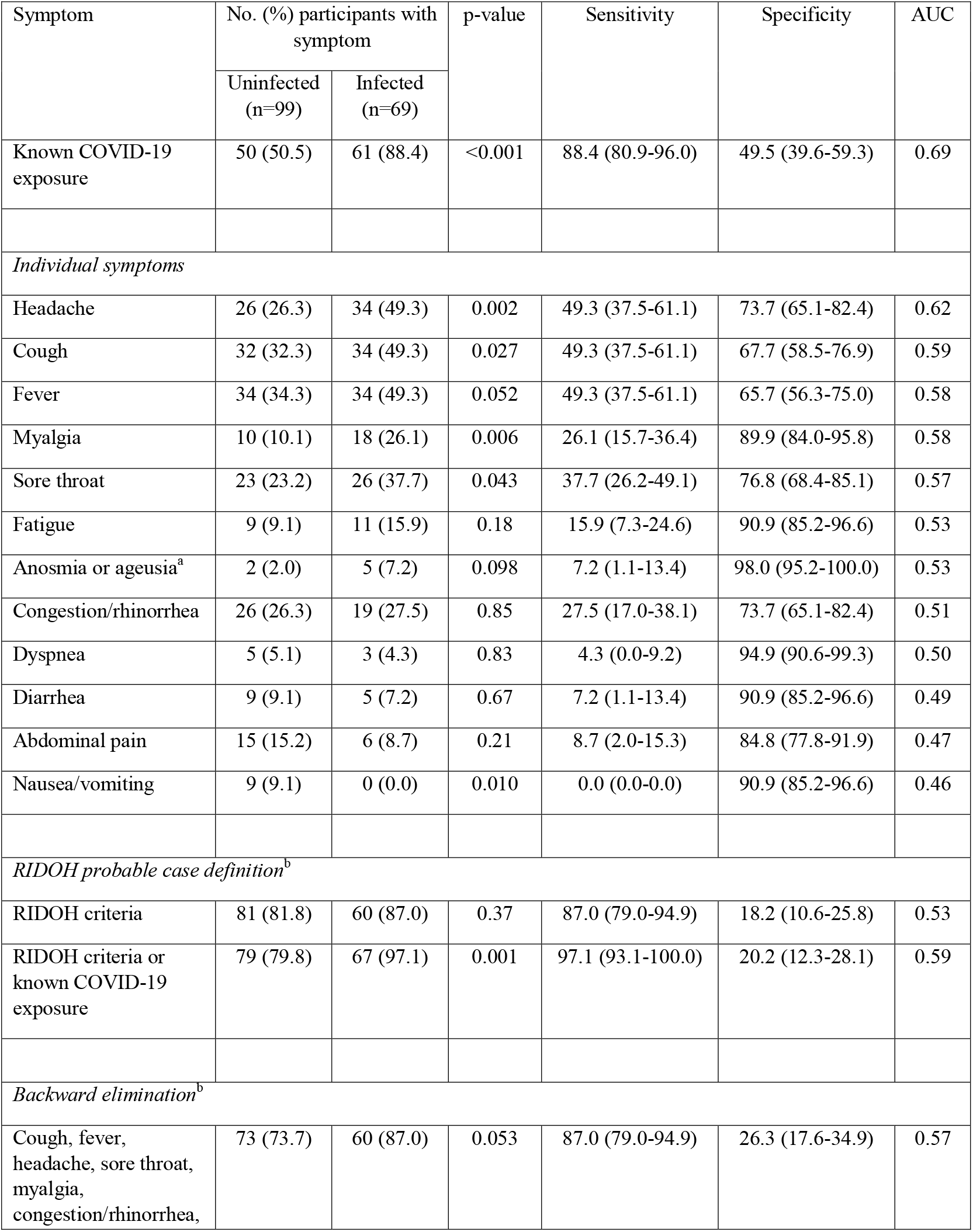
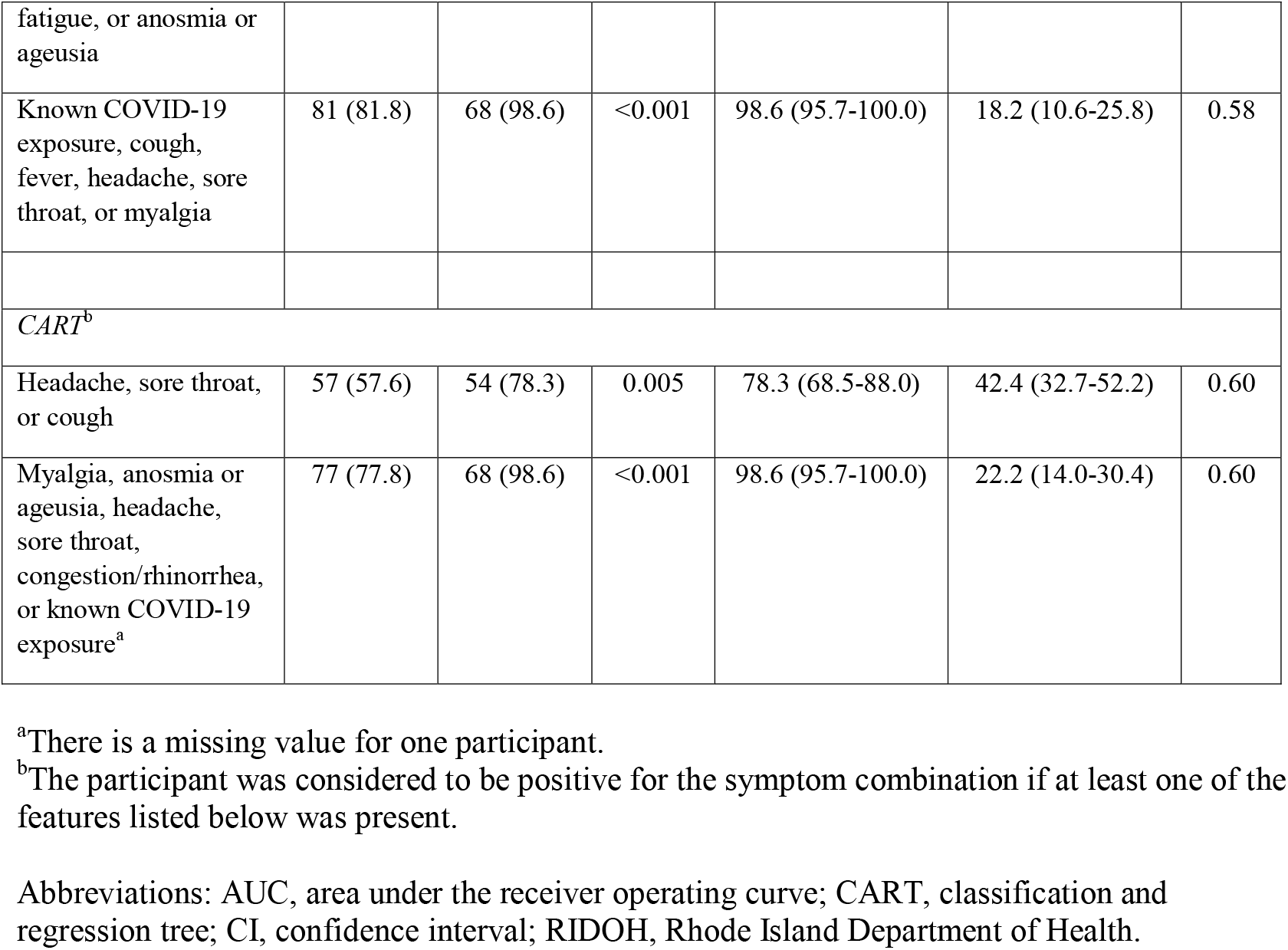
Diagnostic Value of Symptom Screening in Children 5-11 Years of Age

**Table 5:**
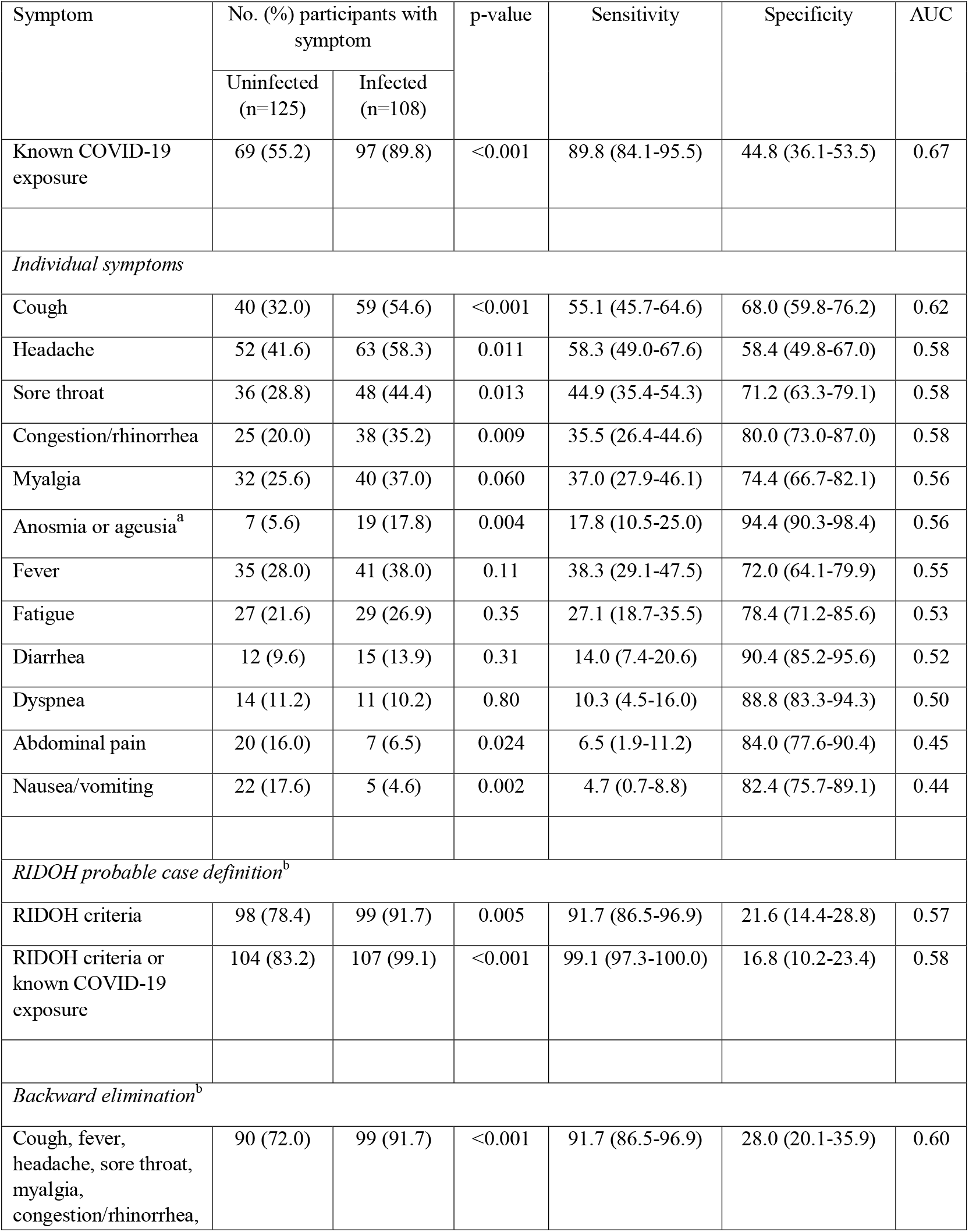
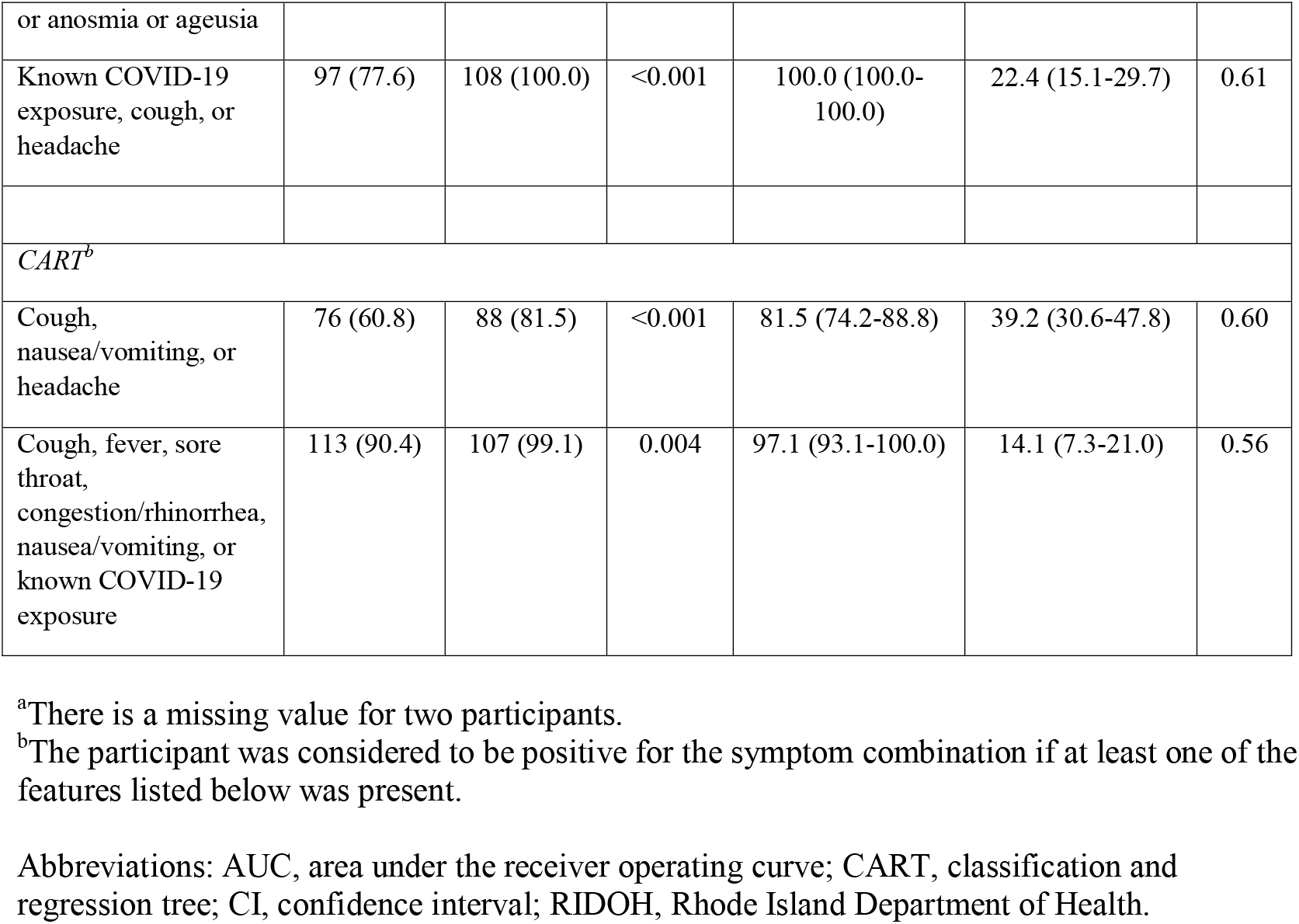
Diagnostic Value of Symptom Screening in Adolescents 12-17 Years of Age

In the sensitivity analysis of participants evaluated with the standardized template before PCR testing, the AUCs of exposure history and individual symptoms were similar to those calculated for the entire study population, with differences of ≤0.02 (Supplementarytable 1 Material).

## Discussion

In this study, we assessed the diagnostic properties of symptoms for SARS-CoV-2 infection in a large pediatric cohort, >90% of whom presented to primary care and were evaluated with a standardized symptom template before PCR testing. We identified symptom combinations with high sensitivity, particularly in conjunction with COVID-19 exposure; however, all symptom combinations had poor specificities. We failed to identify any individual symptom or symptom combination with AUC >0.70, underscoring the importance of widely available SARS-CoV-2 testing with rapid turnaround.

The AUCs observed in our study likely are higher than in the general population for several reasons. First, few study participants were asymptomatic, thus maximizing sensitivity of symptoms. Second, our study took place in the spring and summer; specificities of symptoms are expected to decrease further in the winter as more respiratory viruses circulate. Third, the reliability of COVID-19 exposure history probably was higher in our cohort since many participants were tested during a stay-at-home order. Therefore, they likely had few contacts and were better able to stay informed of the infection status of their contacts.

Reports of pediatric COVID-19 symptoms mostly include hospitalized participants.(3, 14–17) One exception is a study conducted in Alberta, Canada, which used provincial databases to assess the association of symptoms with SARS-CoV-2 PCR positivity.(8) This study found a high positive predictive value for anosmia or ageusia; similarly, we observed a high specificity of these symptoms. Our study differs in a few ways. First, in Alberta, the symptom questionnaire was applied after test results were known, whereas symptoms were assessed before testing in >90% of our cohort, reducing recall bias. Second, we age-stratified participants and detected differences in COVID-19 presentation between age groups. These differences were similar to those reported by the BRAVE study, which evaluated children with a close SARS-CoV-2-infected contact(18): elementary school-aged children were most likely to have asymptomatic COVID-19 (though the difference did not reach statistical significance in our cohort), the youngest children were most likely to be febrile, and adolescents were more likely to report anosmia or ageusia compared to elementary school-aged children.

Our findings have clinical and public health implications. The low AUCs we observed strongly argue against the use of symptoms to diagnose pediatric COVID-19. However, anosmia or ageusia in children ≥5 years old and dyspnea in children 5-11 years old are highly specific, and their presence should alert providers to quickly isolate and test the patient. Exposure history most accurately predicted SARS-CoV-2 infection and should remain a cornerstone of quarantine recommendations. Because of differences between our cohort and daycare and school attendees, the diagnostic characteristics we observed may not be generalizable to that group. However, our findings suggest that different age groups need distinct symptom screening criteria. Additionally, because of the low specificities of most symptoms, easily accessible tests with rapid turnaround times are critical to minimize unnecessary absences.

With respect to the secondary aim of our study, we identified Hispanic ethnicity as an independent risk factor for COVID-19 compared to the reference group of NH Black. Both the BRAVE study and another study conducted in Washington, DC similarly found significantly higher SARS-CoV-2 positivity in Hispanic children.(18, 19) Further investigation is needed to clarify the contribution of various factors—including multigenerational or multi-family housing, the inability to work from home, and language barriers—to these higher positivity rates.(20–24) The DC study reported that NH Blacks also had higher positivity rates than NH Whites, but we did not detect a difference between these groups, potentially due to insufficient statistical power.

This study had limitations. Data were collected early in the pandemic; however, symptoms are not expected to change over the course of the pandemic, and the clinical and public health implications of this study remain relevant and practical. As previously discussed, the AUCs of exposure and symptoms that we observed may represent a “best case scenario,” but this possibility only strengthens the overarching message that symptoms are poorly predictive of COVID-19.

Despite these limitations, our assessment of the diagnostic value of symptoms fills an important gap in the pediatric COVID-19 literature. The poor AUCs we observed mean that symptoms should not be used alone to identify pediatric SARS-CoV-2 infection, and underscore the importance of widely available and efficient testing.

## Data Availability

Deidentified individual participant data will not be made available.

## Acknowledgements

The authors thank Jennifer Friedman and Philip Chan.

## Conflict of Interest Disclosures (includes financial disclosures)

None declared.

## Data Sharing Statement

Deidentified individual participant data will not be made available.

## Author Contributions

Prof. Weng conceptualized the study, collected the data, interpreted the results, drafted the manuscript, and reviewed the manuscript for content.

Prof. Butt conceptualized the study, collected the data, and reviewed the manuscript for content.

Dr. Brooks conducted the analyses, interpreted the results, and reviewed the manuscript for content.

Dr. Clarke conceptualized the study, collected the data, and reviewed the manuscript for content.

Prof. Jenkins provided input for analyses, interpreted the results, and reviewed the manuscript for content.

Prof. Holland conceptualized the study, and reviewed the manuscript for content.

Prof. Chiang conceptualized the study, conducted the analyses, interpreted the results, drafted the manuscript, and reviewed the manuscript for content.

All authors approved the final manuscript as submitted and agree to be accountable for all aspects of the work.

## Abbreviations

SARS-CoV-2: severe acute respiratory syndrome coronavirus 2
COVID-19: coronavirus disease 2019
RT-PCR: reverse transcriptase polymerase chain reaction
NH: non-Hispanic
BMI: body mass index
CART: combination generated by classification and regression tree
IQR: interquartile range
AUC: area under the receiver operating curve

