## Supplementary Material for "Diagnostic Value of Symptoms for Pediatric SARS-CoV-2 Infection in a Primary Care Setting"

Supplementary Table 1: Differences between Patients Included and Excluded in the Study

|  | Excluded (n=248) | Included (n=555) | Total (n=803) | p-value |
| --- | --- | --- | --- | --- |
| Age category |  |  |  | 0.78 |
| 0-4 years | 71 (28.6%) | 154 (27.7%) | 225 (28.0%) |  |
| 5-11 years | 69 (27.8%) | 168 (30.3%) | 237 (29.5%) |  |
| 12-17 years | 108 (43.5%) | 233 (42.0%) | 341 (42.5%) |  |
| Race/ethnicity |  |  |  | <0.001 |
| Hispanic | 153 (61.7%) | 459 (82.7%) | 612 (76.2%) |  |
| NH Black | 28 (11.3%) | 26 (4.7%) | 54 (6.7%) |  |
| NH White | 15 (6.0%) | 30 (5.4%) | 45 (5.6%) |  |
| NH Other | 52 (21.0%) | 40 (7.2%) | 92 (11.5%) |  |
| Uninsured | 32 (12.9%) | 37 (6.7%) | 69 (8.6%) | 0.004 |
| SARS-CoV-2 PCR-positive | 38 (15.3%) | 217 (39.1%) | 255 (31.8%) | <0.001 |

Of 52 excluded patients in the NH Other category, 5 (9.6%) were Asian; 6 (11.5%), >1 race; 1 (1.9%), Pacific Islander; 40 (76.9%), Unknown. Of 40 included patients in the NH Other category, 9 (22.5%) were Asian; 8 (20.0%), >1 race; 1 (2.5%), Pacific Islander; 22 (55.0%), Unknown.

Supplementary Table 2: Age-Stratified Clinical Presentation among 217 SARS-CoV-2-Positive Participants

|  | Total (n=217) | | 0-4 years (n=40) | | 5-11 years (n=69) | | 12-17 years (n=108) | | *P*-value |
| --- | --- | --- | --- | --- | --- | --- | --- | --- | --- |
|  | No. | % (95% CI) | No. | % (95% CI) | No. | % (95% CI) | No. | % (95% CI) |  |
| No symptoms | 20 | 9.2 (5.4, 13.1) | 2 | 5.0 (0.0, 11.8) | 9 | 13.0 (5.1, 21.0) | 9 | 8.3 (3.1, 13.5) | 0.34 |
| Fever | 103 | 47.5 (40.8, 54.1) | 28 | 70.0 (55.8, 84.2) | 34 | 49.3 (37.5, 61.1) | 41 | 38.0 (28.8, 47.1) | 0.002 |
| Fatigue | 43 | 19.8 (14.5, 25.1) | 3 | 7.5 (0.0, 15.7) | 11 | 15.9 (7.3, 24.6) | 29 | 26.9 (18.5, 35.2) | 0.020 |
| Myalgia | NA |  | NA |  | 18 | 26.1 (15.7, 36.4) | 40 | 37.0 (27.9, 46.1) | 0.13 |
| Headache | NA |  | NA |  | 34 | 49.3 (37.5, 61.1) | 63 | 58.3 (49.0, 67.6) | 0.24 |
| Cough | 119 | 54.8 (48.2, 61.5) | 26 | 65.0 (50.2, 79.8) | 34 | 49.3 (37.5, 61.1) | 59 | 54.6 (45.2, 64.0) | 0.28 |
| Dyspnea | 21 | 9.7 (5.7, 13.6) | 7 | 17.5 (5.7, 29.3) | 3 | 4.3 (0.0, 9.2) | 11 | 10.2 (4.5, 15.9) | 0.079 |
| Sore throat | NA |  | NA |  | 26 | 37.7 (26.2, 49.1) | 48 | 44.4 (35.1, 53.8) | 0.37 |
| Congestion/rhinorrhea | 72 | 33.2 (26.9, 39.4) | 15 | 37.5 (22.5, 52.5) | 19 | 27.5 (17.0, 38.1) | 38 | 35.2 (26.2, 44.2) | 0.47 |
| Anosmia/ageusia | NA |  | NA |  | 5 | 7.2 (1.1, 13.4) | 19 | 17.6 (10.4, 24.8) | 0.047 |
| Abdominal pain | NA |  | NA |  | 6 | 8.7 (2.0, 15.3) | 7 | 6.5 (1.8, 11.1) | 0.58 |
| Nausea/vomiting | 8 | 3.7 (1.2, 6.2) | 3 | 7.6 (0.0, 15.7) | 0 | 0.0 (0.0, 0.0) | 5 | 4.6 (0.7, 8.6) | 0.098 |
| Diarrhea | 28 | 12.9 (8.4, 17.4) | 8 | 20.0 (7.6, 32.4) | 5 | 7.2 (1.1, 13.4) | 15 | 13.9 (7.4, 20.4) | 0.15 |

Myalgia, headache, sore throat, abdominal pain, nausea, and anosmia/ageusia were not assessed in children 0-4 years-old due to the lower reliability of these symptoms in this group.

Abbreviations: CI, confidence interval; HEENT, head/eyes/ears/nose/throat; NA, not applicable.

Supplementary Table 3: Backward Elimination, Children 0-4 Years of Age, Symptoms Only

| Symptom(s) removed | No. (%) participants with symptom | | p-value | Sensitivity  (95% CI) | Specificity  (95% CI) | AUC |
| --- | --- | --- | --- | --- | --- | --- |
|  | Uninfected (n=115) | Infected (n=40) |  |  |  |  |
| None | 88 (77.2) | 38 (95.0) | 0.012 | 95.0 (88.2-100.0) | 22.8 (15.1-30.5) | 0.59 |
| Vomiting | 87 (76.3) | 38 (95.0) | 0.009 | 95.0 (88.2-100.0) | 23.7 (15.9-31.5) | 0.59 |
| Vomiting + fatigue | 87 (76.3) | 38 (95.0) | 0.009 | 95.0 (88.2-100.0) | 23.7 (15.9-31.5) | 0.59 |
| Vomiting + fatigue + diarrhea | 83 (72.8) | 37 (92.5) | 0.010 | 92.5 (84.3-100.0) | 27.2 (19.0-35.4) | 0.60 |

Abbreviations: AUC, area under the receiver operating curve; CI, confidence interval.

Supplementary Table 4: Backward Elimination, Children 0-4 Years of Age, Symptoms and Exposure

| Symptom(s) removed | No. (%) participants with symptom | | p-value | Sensitivity  (95% CI) | Specificity  (95% CI) | AUC |
| --- | --- | --- | --- | --- | --- | --- |
|  | Uninfected (n=115) | Infected (n=40) |  |  |  |  |
| None | 99 (86.8) | 40 (100.0) | 0.016 | 100.0 (100.0-100.0) | 13.2 (7.0-19.4) | 0.57 |
| Vomiting | 98 (86.0) | 40 (100.0) | 0.012 | 100.0 (100.0-100.0) | 14.0 (7.7-20.4) | 0.57 |
| Vomiting + fatigue | 98 (86.0) | 40 (100.0) | 0.012 | 100.0 (100.0-100.0) | 14.0 (7.7-20.4) | 0.57 |
| Vomiting + fatigue + diarrhea | 96 (84.2) | 39 (97.5) | 0.028 | 97.5 (92.7-100.0) | 15.8 (9.1-22.5) | 0.57 |

Abbreviations: AUC, area under the receiver operating curve; CI, confidence interval.

Supplementary Table 5: Backward Elimination, Children 6-11 Years of Age, Symptoms Only

| Symptom(s) removed | No. (%) participants with symptom | | p-value | Sensitivity  (95% CI) | Specificity  (95% CI) | AUC |
| --- | --- | --- | --- | --- | --- | --- |
|  | Uninfected (n=99) | Infected (n=69) |  |  |  |  |
| None | 81 (81.8) | 60 (87.0) | 0.37 | 87.0 (79.0-94.9) | 18.2 (10.6-25.8) | 0.53 |
| Nausea/vomiting | 78 (78.8) | 60 (87.0) | 0.17 | 87.0 (79.0-94.9) | 21.2 (13.2-29.3) | 0.54 |
| Nausea/vomiting + abdominal pain | 77 (77.8) | 60 (87.0) | 0.13 | 87.0 (79.0-94.9) | 22.2 (14.0-30.4) | 0.55 |
| Nausea/vomiting + abdominal pain + diarrhea | 74 (74.7) | 60 (87.0) | 0.053 | 87.0 (79.0-94.9) | 25.3 (16.7-33.8) | 0.56 |
| Nausea/vomiting + abdominal pain + diarrhea + dyspnea | 73 (73.7) | 60 (87.0) | 0.053 | 87.0 (79.0-94.9) | 26.3 (17.6-34.9) | 0.57 |
| Nausea/vomiting + abdominal pain + diarrhea + dyspnea + congestion/rhinorrhea | 68 (68.7) | 59 (85.5) | 0.013 | 85.5 (77.2-93.8) | 31.3 (22.2-40.4) | 0.58 |

Abbreviations: AUC, area under the receiver operating curve; CI, confidence interval.

Supplementary Table 6: Backward Elimination, Children 6-11 Years of Age, Symptoms and Exposure

| Symptom(s) removed | No. (%) participants with symptom | | p-value | Sensitivity  (95% CI) | Specificity  (95% CI) | AUC |
| --- | --- | --- | --- | --- | --- | --- |
|  | Uninfected (n=99) | Infected (n=69) |  |  |  |  |
| None | 92 (92.9) | 68 (98.6) | 0.092 | 98.6 (95.7-100.0) | 7.1 (2.0-12.1) | 0.53 |
| Nausea/vomiting | 89 (89.9) | 68 (98.6) | 0.026 | 98.6 (95.7-100.0) | 10.1 (4.2-16.0) | 0.54 |
| Nausea/vomiting + abdominal pain | 88 (88.9) | 68 (98.6) | 0.017 | 98.6 (95.7-100.0) | 11.1 (4.9-17.3) | 0.55 |
| Nausea/vomiting + abdominal pain + diarrhea | 85 (85.9) | 68 (98.6) | 0.005 | 98.6 (95.7-100.0) | 14.1 (7.3-21.0) | 0.56 |
| Nausea/vomiting + abdominal pain + diarrhea + dyspnea | 84 (84.8) | 68 (98.6) | 0.005 | 98.6 (95.7-100.0) | 15.2 (8.1-22.2) | 0.57 |
| Nausea/vomiting + abdominal pain + diarrhea + shortness of breath + congestion/rhinorrhea | 82 (82.8) | 68 (98.6) | 0.001 | 98.6 (95.7-100.0) | 17.2 (9.7-24.6) | 0.58 |
| Nausea/vomiting + abdominal pain + diarrhea + dyspnea + congestion/rhinorrhea + anosmia/ageusia | 82 (82.8) | 68 (98.6) | <0.001 | 98.6 (95.7-100.0) | 17.2 (9.7-24.6) | 0.58 |
| Nausea/vomiting + abdominal pain + diarrhea + dyspnea + congestion/rhinorrhea + anosmia/ageusia + fatigue | 81 (81.8) | 68 (98.6) | <0.001 | 98.6 (95.7-100.0) | 18.2 (10.6-25.8) | 0.58 |
| Nausea/vomiting + abdominal pain + diarrhea + dyspnea + congestion/rhinorrhea + anosmia/ageusia + fatigue + sore throat | 79 (79.8) | 67 (97.1) | 0.001 | 97.1 (93.1-100.0) | 20.2 (12.3-28.1) | 0.59 |

Abbreviations: AUC, area under the receiver operating curve; CI, confidence interval.

Supplementary Table 7: Backward Elimination, Children 12-17 Years of Age, Symptoms Only

| Symptom(s) removed | No. (%) participants with symptom | | p-value | Sensitivity  (95% CI) | Specificity  (95% CI) | AUC |
| --- | --- | --- | --- | --- | --- | --- |
|  | Uninfected (n=125) | Infected (n=108) |  |  |  |  |
| None | 99 (79.2) | 99 (91.7) | 0.008 | 91.7 (86.5-96.9) | 20.8 (13.7-27.9) | 0.56 |
| Nausea/vomiting | 99 (79.2) | 99 (91.7) | 0.008 | 91.7 (86.5-96.9) | 20.8 (13.7-27.9) | 0.56 |
| Nausea/vomiting + abdominal pain | 94 (75.2) | 99 (91.7) | <0.001 | 91.7 (86.5-96.9) | 24.8 (17.2-32.4) | 0.58 |
| Nausea/vomiting + abdominal pain + dyspnea | 94 (75.2) | 99 (91.7) | <0.001 | 91.7 (86.5-96.9) | 24.8 (17.2-32.4) | 0.58 |
| Nausea/vomiting + abdominal pain + dyspnea + diarrhea | 92 (73.6) | 99 (91.7) | <0.001 | 91.7 (86.5-96.9) | 26.4 (18.7-34.1) | 0.59 |
| Nausea/vomiting + abdominal pain + dyspnea + diarrhea + fatigue | 90 (72.0) | 99 (91.7) | <0.001 | 91.7 (86.5-96.9) | 28.0 (20.1-35.9) | 0.60 |
| Nausea/vomiting + abdominal pain + dyspnea + diarrhea + fatigue + fever* | 87 (69.6) | 98 (90.7) | <0.001 | 90.7 (85.3-96.2) | 30.4 (22.3-38.5) | 0.61 |

Abbreviations: AUC, area under the receiver operating curve; CI, confidence interval.

Supplementary Table 8: Backward Elimination, Children 12-17 Years of Age, Symptoms and Exposure

| Symptom(s) removed | No. (%) participants with symptom | | p-value | Sensitivity  (95% CI) | Specificity  (95% CI) | AUC |
| --- | --- | --- | --- | --- | --- | --- |
|  | Uninfected (n=125) | Infected (n=108) |  |  |  |  |
| None | 115 (92.0) | 108 (100.0) | 0.003 | 100.0 (100.0-100.0) | 8.0 (3.2-12.8) | 0.54 |
| Nausea/vomiting | 115 (92.0) | 108 (100.0) | 0.003 | 100.0 (100.0-100.0) | 8.0 (3.2-12.8) | 0.54 |
| Nausea/vomiting + abdominal pain | 110 (88.0) | 108 (100.0) | <0.001 | 100.0 (100.0-100.0) | 12.0 (6.3-17.7) | 0.56 |
| Nausea/vomiting + abdominal pain + dyspnea | 110 (88.0) | 108 (100.0) | <0.001 | 100.0 (100.0-100.0) | 12.0 (6.3-17.7) | 0.56 |
| Nausea/vomiting + abdominal pain + dyspnea + diarrhea | 108 (86.4) | 108 (100.0) | <0.001 | 100.0 (100.0-100.0) | 13.6 (7.6-19.6) | 0.57 |
| Nausea/vomiting + abdominal pain + dyspnea + diarrhea + fatigue | 107 (85.6) | 108 (100.0) | <0.001 | 100.0 (100.0-100.0) | 14.4 (8.2-20.6) | 0.57 |
| Nausea/vomiting + abdominal pain + dyspnea + diarrhea + fatigue + fever^a^ | 106 (84.8) | 108 (100.0) | <0.001 | 100.0 (100.0-100.0) | 15.2 (8.9-21.5) | 0.58 |
| Nausea/vomiting + abdominal pain + dyspnea + diarrhea + fatigue + fever + anosmia/ageusia | 106 (84.8) | 108 (100.0) | <0.001 | 100.0 (100.0-100.0) | 15.2 (8.9-21.5) | 0.58 |
| Nausea/vomiting + abdominal pain + dyspnea + diarrhea + fatigue + fever + anosmia/ageusia + myalgia | 104 (83.2) | 108 (100.0) | <0.001 | 100.0 (100.0-100.0) | 16.8 (10.2-23.4) | 0.58 |
| Nausea/vomiting + abdominal pain + dyspnea + diarrhea + fatigue + fever + anosmia/ageusia + myalgia + congestion/rhinorrhea | 102 (81.6) | 108 (100.0) | <0.001 | 100.0 (100.0-100.0) | 18.4 (11.6-25.2) | 0.59 |
| Nausea/vomiting + abdominal pain + dyspnea + diarrhea + fatigue + fever + anosmia/ageusia + myalgia + congestion/rhinorrhea  + sore throat | 97 (77.6) | 108 (100.0) | <0.001 | 100.0 (100.0-100.0) | 22.4 (15.1-29.7) | 0.61 |
| Nausea/vomiting + abdominal pain + dyspnea + diarrhea + fatigue + fever + anosmia/ageusia + myalgia + congestion/rhinorrhea  + sore throat + headache | 82 (65.6) | 106 (98.1) | <0.001 | 98.1 (95.6-100.0) | 34.4 (26.1-42.7) | 0.66 |

^a^There is a missing value for one participant.

Abbreviations: AUC, area under the receiver operating curve; CI, confidence interval.

Supplementary Table 9: Sensitivity Analysis of Diagnostic Value of Exposure and Individual Symptoms in Children 0-4 Years of Age (n=138)

|  | No. (%) participants with symptom | | p-value | Sensitivity  (95% CI) | Specificity  (95% CI) | AUC |
| --- | --- | --- | --- | --- | --- | --- |
|  | Uninfected (n=96) | Infected (n=37) |  |  |  |  |
| Known COVID-19 exposure | 29 (30.2) | 28 (75.7) | <0.001 | 75.7 (61.9-89.5) | 69.8 (60.6-79.0) | 0.73 |
| *Individual symptoms* | | | | | | |
| Cough | 29 (30.2) | 25 (67.6) | <0.001 | 67.6 (52.5-82.7) | 69.8 (60.6-79.0) | 0.69 |
| Congestion/rhinorrhea | 22 (22.9) | 15 (40.5) | 0.042 | 40.5 (24.7-56.4) | 77.3 (69.0-85.7) | 0.59 |
| Fever | 58 (60.4) | 27 (73.0) | 0.18 | 73.0 (58.7-87.3) | 39.2 (29.5-48.9) | 0.56 |
| Dyspnea | 11 (11.5) | 5 (13.5) | 0.74 | 13.5 (2.5-24.5) | 88.5 (82.2-94.9) | 0.51 |
| Diarrhea | 19 (19.8) | 8 (21.6) | 0.81 | 21.6 (8.4-34.9) | 80.2 (72.2-88.2) | 0.51 |
| Fatigue | 7 (7.3) | 3 (8.1) | 0.87 | 8.1 (0.0-16.9) | 92.7 (87.5-97.9) | 0.50 |
| Vomiting^a^ | 15 (15.6) | 3 (8.3) | 0.28 | 8.3 (0.0-17.4) | 84.4 (77.1-91.6) | 0.46 |

^a^There is a missing value for one participant.

^b^Cough or shortness of breath alone, or two of any of the following: fever, fatigue, congestion/rhinorrhea, nausea/vomiting, and diarrhea.

Abbreviations: AUC, area under the receiver operating curve; CI, confidence interval.

Supplementary Table 10: Sensitivity Analysis of Diagnostic Value of Exposure and Individual Symptoms in Children 5-11 Years of Age

| Symptom | No. (%) participants with symptom | | p-value | Sensitivity | Specificity | AUC |
| --- | --- | --- | --- | --- | --- | --- |
|  | Uninfected (n=91) | Infected (n=67) |  |  |  |  |
| Known COVID-19 exposure | 50 (54.9) | 60 (89.6) | <0.001 | 89.6 (82.2-96.9) | 45.1 (34.8-55.3) | 0.67 |
| *Individual symptoms* | | | | | | |
| Headache | 25 (27.5) | 33 (49.3) | 0.005 | 49.3 (37.3-61.2) | 73.1 (64.1-82.1) | 0.61 |
| Cough | 32 (35.2) | 34 (50.7) | 0.050 | 50.7 (38.8-62.7) | 64.8 (55.0-74.6) | 0.58 |
| Myalgia | 10 (11.0) | 18 (26.9) | 0.010 | 26.9 (16.3-37.5) | 89.0 (82.6-95.4) | 0.58 |
| Fever | 33 (36.3) | 33 (49.3) | 0.102 | 49.3 (37.3-61.2) | 63.7 (53.9-73.6) | 0.57 |
| Sore throat | 23 (25.3) | 26 (38.8) | 0.069 | 38.8 (27.1-50.5) | 74.7 (65.8-83.7) | 0.57 |
| Fatigue | 9 (9.7) | 11 (16.4) | 0.20 | 16.4 (7.5-25.3) | 90.1 (84.0-96.2) | 0.53 |
| Anosmia/ageusia^a^ | 2 (2.2) | 5 (7.5) | 0.12 | 7.5 (1.2-13.8) | 97.8 (94.7-100.0) | 0.53 |
| Congestion/rhinorrhea | 26 (28.6) | 19 (28.4) | 0.98 | 28.4 (17.6-39.2) | 71.4 (62.1-80.7) | 0.50 |
| Dyspnea | 4 (4.4) | 3 (4.5) | 0.98 | 4.5 (0.0-9.4) | 95.6 (91.4-99.8) | 0.50 |
| Diarrhea | 9 (9.9) | 5 (7.5) | 0.60 | 7.5 (1.2-13.8) | 90.1 (84.0-96.2) | 0.49 |
| Abdominal pain | 14 (15.4) | 5 (7.5) | 0.13 | 7.5 (1.2-13.8) | 84.6 (77.2-92.0) | 0.46 |
| Nausea/vomiting | 9 (9.9) | 0 (0.0) | 0.008 | 0.0 (0.0-0.0) | 90.1 (84.0-96.2) | 0.45 |

^a^There is a missing value for one participant.

Abbreviations: AUC, area under the receiver operating curve; CI, confidence interval.

Supplementary Table 11: Sensitivity Analysis of Diagnostic Value of Exposure and Individual Symptoms in Children 12-17 Years of Age

| Symptom | No. (%) participants with symptom | | p-value | Sensitivity | Specificity | AUC |
| --- | --- | --- | --- | --- | --- | --- |
|  | Uninfected (n=114) | Infected (n=106) |  |  |  |  |
| Known COVID-19 exposure | 68 (59.6) | 95 (89.6) | <0.001 | 89.6 (83.8-95.4) | 40.4 (31.3-49.4) | 0.65 |
| *Individual symptoms* | | | | | | |
| Cough | 39 (34.2) | 58 (54.7) | 0.002 | 54.7 (45.2-64.2) | 65.8 (57.1-74.5) | 0.60 |
| Headache | 51 (44.7) | 63 (59.4) | 0.029 | 59.4 (50.1-68.8) | 55.3 (46.1-64.4) | 0.57 |
| Congestion/rhinorrhea | 25 (21.9) | 38 (35.8) | 0.022 | 35.8 (26.7-45.0) | 78.1 (70.5-85.7) | 0.57 |
| Sore throat | 36 (31.6) | 47 (44.3) | 0.051 | 44.3 (34.9-53.8) | 68.4 (59.9-77.0) | 0.56 |
| Myalgia | 32 (28.1) | 40 (37.7) | 0.13 | 37.7 (28.5-47.0) | 71.9 (63.7-80.2) | 0.55 |
| Anosmia/ageusia^a^ | 7 (6.1) | 19 (18.1) | 0.007 | 18.1 (10.7-25.5) | 93.8 (89.4-98.2) | 0.56 |
| Fever | 34 (29.8) | 41 (38.7) | 0.17 | 38.7 (29.4-48.0) | 70.2 (61.8-78.6) | 0.54 |
| Fatigue | 26 (22.8) | 29 (27.4) | 0.44 | 27.4 (18.9-35.8) | 77.2 (69.5-84.9) | 0.52 |
| Diarrhea | 11 (9.6) | 15 (14.2) | 0.30 | 14.2 (7.5-20.8) | 90.4 (84.9-95.8) | 0.52 |
| Dyspnea | 13 (11.4) | 11 (10.4) | 0.81 | 10.4 (4.6-16.2) | 88.6 (82.8-94.4) | 0.50 |
| Abdominal pain | 15 (13.2) | 7 (6.6) | 0.11 | 6.6 (1.9-11.3) | 86.8 (80.6-93.0) | 0.47 |
| Nausea/vomiting | 19 (16.7) | 5 (4.7) | 0.005 | 4.7 (0.1-8.8) | 83.3 (76.5-90.2) | 0.44 |

^a^There is a missing value for two participants.

Abbreviations: AUC, area under the receiver operating curve; CI, confidence interval.
